# Investigating the shared genetic architecture between chronic pain and depression

**DOI:** 10.64898/2026.04.03.26348156

**Authors:** Hannah Casey, Xueyi Shen, Laurence Nisbet, Marie T. Fallon, Daniel J. Smith, Rona J. Strawbridge, Heather C. Whalley

**Affiliations:** Division of Psychiatry, University of Edinburgh, Edinburgh, UK; Department of Palliative Medicine, Edinburgh Cancer Research Centre, University of Edinburgh, Edinburgh, UK; Institute of Health and Wellbeing, University of Glasgow, Glasgow, UK; Cardiovascular Medicine Unit, Department of Medicine, Karolinska Institute, Stockholm, Sweden; Generation Scotland, Institute for Genetics and Cancer, University of Edinburgh, Edinburgh, UK

**Keywords:** Chronic pain, depression, comorbidity, genetic overlap, genetic colocalisation, gene expression

## Abstract

**Background:** Chronic pain and depression are common disorders and leading causes of disability worldwide. They frequently co-occur and show substantial genetic correlation, indicating a shared genetic basis. However, the locus-specific architecture of this overlap remains poorly characterised and may yield important insights into the pathophysiology of their comorbidity.

**Methods:** Using the largest currently available European-ancestry genome-wide association studies of major depressive disorder (MDD) (n = 1,639,572) and multisite chronic pain (MCP) (n = 387,649), we estimated the polygenic overlap between traits using the bivariate causal mixture model (MiXeR), identified shared loci via conjunctional false discovery rate (conjFDR), and tested colocalisation with each trait and genetically regulated gene expression in 13 brain tissues.

**Results:** MiXeR analysis demonstrated a high degree of directionally consistent polygenic overlap between MDD and MCP. Subsequent conjFDR analysis identified 375 shared loci, 22 of which showed cross-trait colocalisation between the MDD and MCP signals. Gene mapping and enrichment of shared loci implicated several biological processes, including cadherin-mediated cell-cell adhesion and translational initiation. Gene expression colocalisation in brain tissue highlighted *protein phosphatase 6 catalytic subunit* (*PPP6C*) and *suppressor of cancer cell invasion* (*SCAI*) in both disorders.

**Conclusion:** Overall, these findings have enhanced our understanding of the complex relationship between chronic pain and depression by identifying potential shared molecular mechanisms that warrant further study as targets for prevention and treatment.

## Introduction

The frequent co-occurrence of chronic pain and depression is well established; a recent meta-analysis study of 376 studies across 50 countries estimates that 39.3% of adults with chronic pain have clinical symptoms of depression (Aaron et al., 2025). Individually, chronic pain and depression are leading global causes of disability, with low back pain and depressive disorders identified in the Global Burden of Disease Study 2023 as the top contributors to years lived with disability (YLDs) (Hay et al., 2025). When both disorders co-occur, they are associated with greater symptom severity and functional impairment (Bair et al., 2003) and fewer functional benefits from antidepressant use (Roughan et al., 2021), emphasising the importance of furthering our current understanding of how this comorbidity occurs and can be treated.

Genome-wide association studies (GWAS) have identified the polygenic basis of chronic pain and depression. A recent trans-ancestry GWAS of major depressive disorder (MDD) conducted by the Psychiatric Genomics Consortium (PGC) identified 697 variants and 308 genes associated with MDD, implicating postsynaptic density and neuronal dysregulation (Adams et al., 2025). GWAS of multisite chronic pain (MCP) and specific pain disorders/sites, such as osteoarthritis and chronic back pain, have identified numerous risk variants with pathway enrichment in nervous system development, synaptic/signalling processes and inflammation (Hatzikotoulas et al., 2025; Johnston et al., 2019a; Stanaway et al., 2025). Efforts to understand pleiotropy between the two disorders have also identified substantial genetic correlation between MCP and MDD (r_g_ = 0.53) (Johnston et al., 2019a). Additionally, analyses based on earlier, lower-powered GWASes using a conjunctional false-discovery rate (conjFDR) framework that leverages pleiotropy reported suggestive shared loci at *leucine rich repeat and fibronectin type III domain containing 5* (*LRFN5)* (Johnston et al., 2019b).

While previous studies have provided important evidence of genetic pleiotropy between chronic pain and depression, there are limitations to their methodology that can be addressed by leveraging recent, well-powered GWASes and more robust genetic analysis approaches. Specifically, polygenic overlap between MCP and MDD has previously been quantified as genetic correlation using linkage disequilibrium score regression (LDSC) (Johnston et al., 2019a), which underestimates the extent of genetic overlap in cases of mixed directions of effect across shared genetic loci. To help address this, Frei et al. introduced the bivariate causal mixture model (MiXeR), which quantifies the extent of shared polygenicity by estimating the number of trait-influencing variants in each trait and their overlap, irrespective of effect direction (Frei et al., 2019). Furthermore, leveraging the PGC’s largest-to-date GWAS of MDD alongside well-powered GWAS of MCP to localise shared loci increases statistical power to detect genes jointly implicated in chronic pain and depression (Adams et al., 2025; Johnston et al., 2019a). In addition, genetic colocalisation of putatively shared loci would strengthen evidence for a shared causal variant and extending colocalisation to brain-tissue expression quantitative trait locus (eQTL) would prioritise likely effector genes and tissues, improving mechanistic interpretation.

Therefore, the present study had three main objectives: 1) to quantify the polygenic overlap between MCP and MDD using MiXeR applied to large-scale GWAS summary statistics, 2) to identify shared loci with conjFDR and corroborate shared causal variants with genetic colocalisation; and 3) to test whether brain-tissue eQTLs colocalise with both traits. I also discuss the research and clinical implications of these findings.

## Methods

### GWAS data

We used European-ancestry GWAS summary statistics for MDD from the recent PGC meta-analysis (Adams et al., 2025) to ensure compatibility in linkage disequilibrium (LD) patterns, as ancestry-mismatched LD can bias conjFDR analyses. To prevent sample overlap with the MCP GWAS, all UK Biobank (UKB) participants were excluded. While MiXeR can model overlapping samples, independence is required for the condFDR and conjFDR analyses because overlap can inflate apparent cross-trait enrichment. We also excluded 23andMe cohorts due to data-use restrictions. The final dataset comprised 357,636 MDD cases and 1,281,936 controls.

We used UKB summary statistics for MCP (Johnston et al., 2019a). MCP was defined as the count of anatomical sites (0–7) with pain lasting at least three months; individuals reporting “pain all over the body” were excluded. The analysis included 387,649 UKB participants of self-reported White British ancestry.

### Univariate and bivariate MiXeR analyses

To assess the overlapping genetic architecture between MDD and MCP, we applied MiXeR (v1.3, https://github.com/precimed/mixer). First, the univariate MiXeR model estimated heritability, polygenicity, and discoverability for each trait separately (Holland et al., 2020). In MiXeR, heritability (h^2^_SNP_) is the total variance in a trait attributable to causal variants (non-zero effect); polygenicity is the causal variants count explaining 90% of h^2^_SNP_, and discoverability is the average variance of non-zero effects, reflecting how readily genetic signals can be detected at a given sample size.

The bivariate MiXeR model was used to estimate the additive genetic effects as a mixture of four Gaussian components: variants affecting neither trait, only one of the traits, or both traits (Frei et al., 2019). From this, polygenic overlap was quantified by a Dice coefficient (shared / total causal variants). This reflects the overlap in trait-influencing variants between MDD and MCP. The proportion of shared causal variants with concordant effects (the same effect directions) was also measured. Variants in the major histocompatibility complex (MHC; chr6: 25–35 Mb) were excluded to avoid confounding from complex LD. The 1000 Genome Project phase 3 European subsample was used as an LD reference (Auton et al., 2015).

Model fit was evaluated with the Akaike Information Criterion (AIC). For univariate analyses, a positive AIC indicates the GWAS has sufficient power to justify MiXeR’s mixture model over a simpler LDSC baseline. For bivariate analyses, a positive AIC shows that the fitted overlap is preferred over boundary models assuming minimal (AIC_best vs. min_) or maximal (AIC_best vs. max_) possible overlap.

### Conjunctional FDR analyses

To identify shared genetic variants between MDD and MCP, we applied pleiotropy-informed conjFDR framework using the “pleiofdr” software (https://github.com/precimed/pleiofdr) (Andreassen et al., 2013; Smeland et al., 2020). The method builds on conditional FDR (condFDR), which re-ranks genetic variant test statistics for a primary trait according to their evidence in a secondary trait, thereby boosting power to detect associations. Cross-trait enrichment was visualised with conditional Q-Q plots, which display the distribution of primary-trait P-values within genetic variant strata defined by their association with the secondary trait (p < 0.1, p < 0.01, and p < 0.001). Increasing leftward deflection from the null line with more stringent strata indicates enrichment. To improve model fit, variants within the MHC (chr6: 25–35 Mb) and 8p23.1 (chr8: 7.2–12.5 Mb) regions were excluded before fitting the condFDR model. This exclusion was performed as both genomic regions are comprised of highly correlated variants within a long-range LD region, which have previously been shown to associate with psychiatric and neurological disorders (Giner-Delgado et al., 2019; Salm et al., 2012). Subsequent conjFDR analysis was then performed to identify genetic variants jointly associated with MDD and MCP, with the conjFDR value defined as the maximum of the two mutual condFDR values. The conjFDR approach estimates a conservative posterior probability, indicating the likelihood that a variant is null for either one or both phenotypes, given that the p-values for both phenotypes are lower than the observed p-values. The significance threshold was set as conjFDR < 0.05, as recommended in previous studies (Hindley et al., 2022; Liu et al., 2024).

### Genomic locus definition

Independent loci were defined from the conjFDR results using the “sumstats.py” script from precimed (https://github.com/precimed/python_convert), which follows the FUMA SNP2GENE protocol (Watanabe et al., 2017). “Independent SNPs” (single nucleotide polymorphisms) were those with conjFDR < 0.05 and pairwise LD r^2^ < 0.6. From these, a subset of “lead SNPs” was derived by further clumping at LD r^2^ < 0.1. For each independent SNP, we defined “candidate SNPs” as all variants in LD r^2^ ≥ 0.6; locus boundaries were taken as the outermost positions of these candidate SNPs. When the boundaries of two loci lay within 250 kb of one another, the loci were merged into a single region. LD was estimated using the 1000 Genomes Project Phase 3 European reference panel (Auton et al., 2015).

### Cross-trait genetic colocalisation analysis

To test whether MDD and MCP share the same causal variant at loci identified by the conjFDR analysis, we performed Bayesian colocalisation of the two traits within each shared locus. Fine-mapping was carried out with Sum of Single Effects (SuSiE) via the COLOC R package to identify credible sets (CS) for each trait at each locus (Wallace, 2021). A CS is a set of variants with an estimated ≥95% probability of containing a causal variant, and SuSiE permits multiple CS within a region. SNP–SNP signed LD correlation (r) matrices were computed from the 1000 Genomes Project Phase 3 European reference panel (Auton et al., 2015). We then performed pairwise colocalisation between CSs from the two traits using COLOC’s default priors for each genetic variant being associated with trait 1 only (P1 = 1×10^−4^), trait 2 only (P2 = 1×10^−4^), or both traits (P12 = 1×10^−5^). Colocalisation was summarised by the posterior probability for hypothesis 4 (PP4), i.e., the probability that both traits share a causal variant. When SuSiE did not identify CS in one or both traits, we ran coloc.abf() without fine-mapping under the single causal variant assumption (Giambartolomei et al., 2014). Strong evidence for colocalisation was defined as PP4 > 0.8.

### Gene prioritisation and functional annotation

We mapped the likely causal variants in each loci found to be colocalised in MDD and MCP to candidate genes using the Locus-to-Gene (L2G) pipeline (Ghoussaini et al., 2021). For each colocalised region, using L2G, genetic variants from the 95% credible sets identified by coloc-SuSiE and the likely causal variant from coloc.abf were linked to their most likely causative genes. L2G integrates multiple lines of variant-to-gene evidence in a supervised model trained on gold-standard gene-trait pairs. Variant features are aggregated across fine-mapped credible sets using posterior probabilities to weight evidence linking each variant to nearby genes. Evidence types include (i) distance from transcript start site (TSS), (ii) molecular quantitative trait loci (QTL) evidence (eQTLs and pQTLs across tissues), (iii) cis-regulatory/chromatin interaction data (e.g., promoter-capture Hi-C and enhancer-promoter links), and (iv) predicted variant consequences/pathogenicity from Ensembl VEP and related annotations (McLaren et al., 2016).

Genes assigned by L2G were annotated using Metascape (v3.5.20250701) (Zhou et al., 2019). Annotations included (i) disease associations from DisGeNET and GeDiPNet; (ii) pathogenic loss-of-function from ClinVar; (iii) prior GWAS associations from the NHGRI-EBI GWAS Catalog; (iv) drug-target annotations from DrugBank; (v) protein function and expression features from the Human Protein Atlas; and (vi) pathway membership and enrichment from Gene Ontology (GO) and Kyoto Encyclopaedia of Genes and Genomes (KEGG). Furthermore, pathway enrichment for GO terms and KEGG pathways was also performed using Metascape. This analysis was performed using default parameters (minimum overlap = 3, enrichment factor > 1.5, P < 0.01).

### Genetic colocalisation with gene expression in brain tissue

Using GTEx v8 brain-tissue eQTL data (European subset), we tested whether our prioritised genes showed cis-eQTL colocalisation with GWAS signals for MCP and MDD, applying the same Bayesian framework described above. For each gene, we considered all variants spanning the gene body extended by 500 kb. Analyses were run separately in GTEx v8 brain tissues: amygdala, anterior cingulate cortex (BA24), caudate (basal ganglia), cerebellum, cerebellar hemisphere, cortex, frontal cortex (BA9), hippocampus, hypothalamus, nucleus accumbens (basal ganglia), putamen (basal ganglia), spinal cord (cervical c-1) and substantia nigra.

## Results

### Genetic overlap between MCP and MDD

Univariate MiXeR was used to estimate the heritability, polygenicity, and discoverability of MDD and MCP (Holland et al., 2020). Both univariate models showed a good model fit, as indicated by positive AIC values. As shown in Figure 1(A) and Supplementary Table 1, both traits were found to be modestly heritable (MDD = 0.051, MCP = 0.072) and highly polygenic, as measured by trait-influencing variants (MDD = 11,139, MCP = 12,537), with approximately 24% lower discoverability in MDD (7.10E-06) than MCP (8.81E-06).

**Figure 1.**
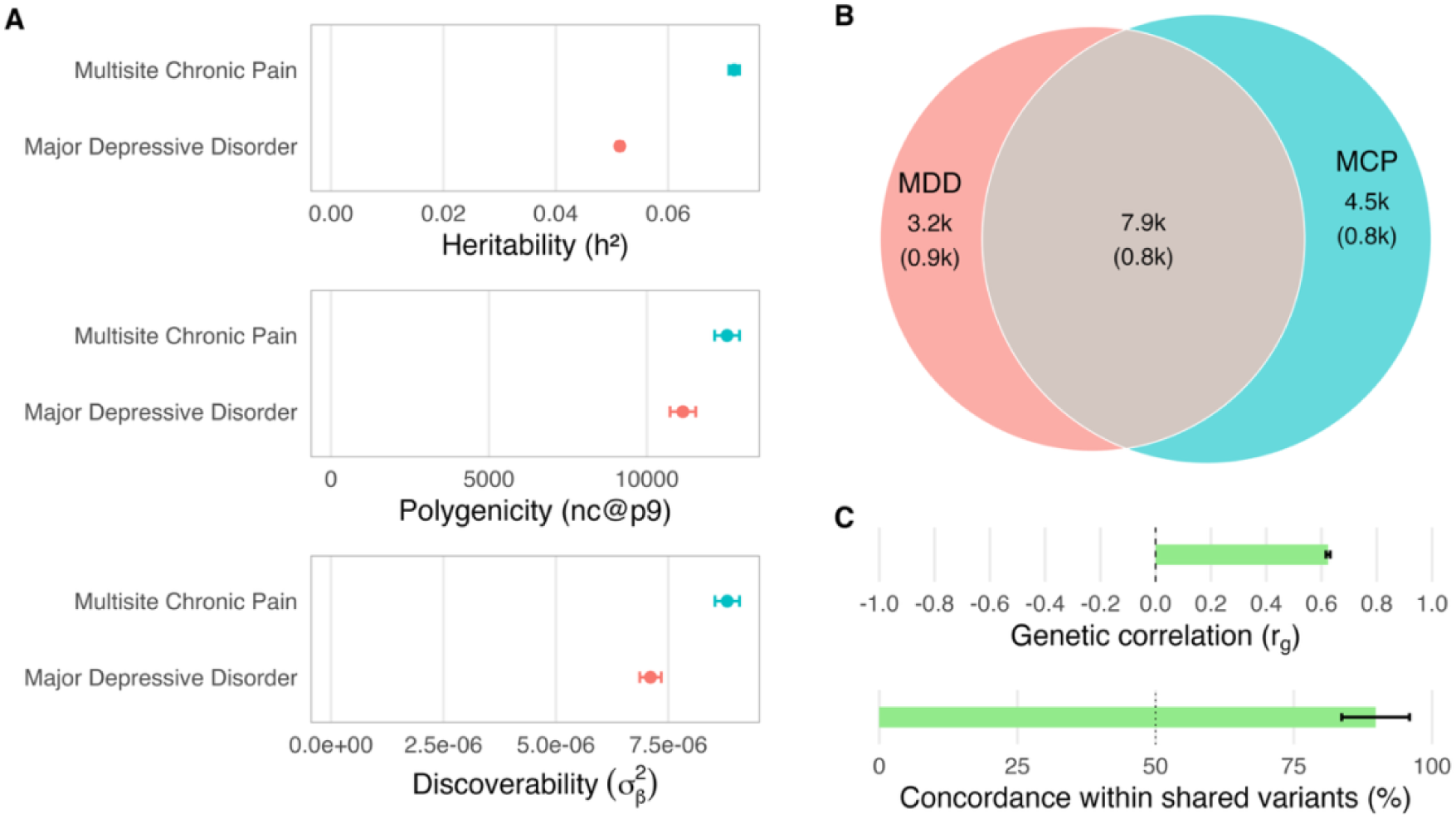
Genetic overlap between major depressive disorder and multisite chronic pain using MiXeR. **A** The heritability, polygenicity, and discoverability of MDD and MCP estimated by the univariate MiXeR model. Error bars represent standard error (SE). **B** Venn diagram showing the estimated overlap and SE of variants between MDD and MCP estimated by the bivariate MiXeR model. The size of each circle represents the extent of each trait’s polygenicity. **C** The genetic correlation and concordance within shared variants between MDD and MCP estimated by the univariate MiXeR model. Error bars represent SE. MCP = multisite chronic pain, MDD = major depressive disorder.

In bivariate MiXeR analysis (Frei et al., 2019), a significant proportion of shared variants was identified between MDD and MCP. When model fit was assessed, a positive AIC_best vs. max_ (1.685) indicated that the bivariate MiXeR model, which allows for trait-specific variation, was a better fit than a model that assumes that all variants of the less polygenic trait are a subset of the more polygenic trait. Conversely, the negative AIC_best vs. min_ (−0.060) indicated the bivariate MiXeR model offered no improvement over the minimum-overlap model defined by the genetic correlation, suggesting little evidence for overlap beyond that implied by r_g_.

The bivariate MiXeR model estimated a large overlap in the causal variants between MDD and MCP (N = 7,942), accounting for 71.3% of the trait-influencing variants of MDD and 63.9% of the trait-influencing variants of MCP (Figure 1(B)); the Dice coefficient quantifying this overlap was 67.4%. The bivariate model also indicated that, among the variants shared between MDD and MCP, the proportion with concordant allelic effects was 89.8%, and the genome-wide cross-trait r_g_ was 0.624 (Figure 1(C)).

The conditional Q-Q plots show cross-trait enrichment between MDD and MCP, consistent with polygenic overlap (Figure 2). In these plots, the blue line shows the p-values of all genetic variants in the primary trait. The orange, green, and red curves are restricted to variants increasingly associated with the conditioning trait (p ≤ 0.100, 0.010, and 0.001, respectively). As the conditioning threshold becomes more stringent, the curves deflect further left from the null diagonal, indicating pleiotropic enrichment of genetic variant associations shared by MDD and MCP.

**Figure 2.**
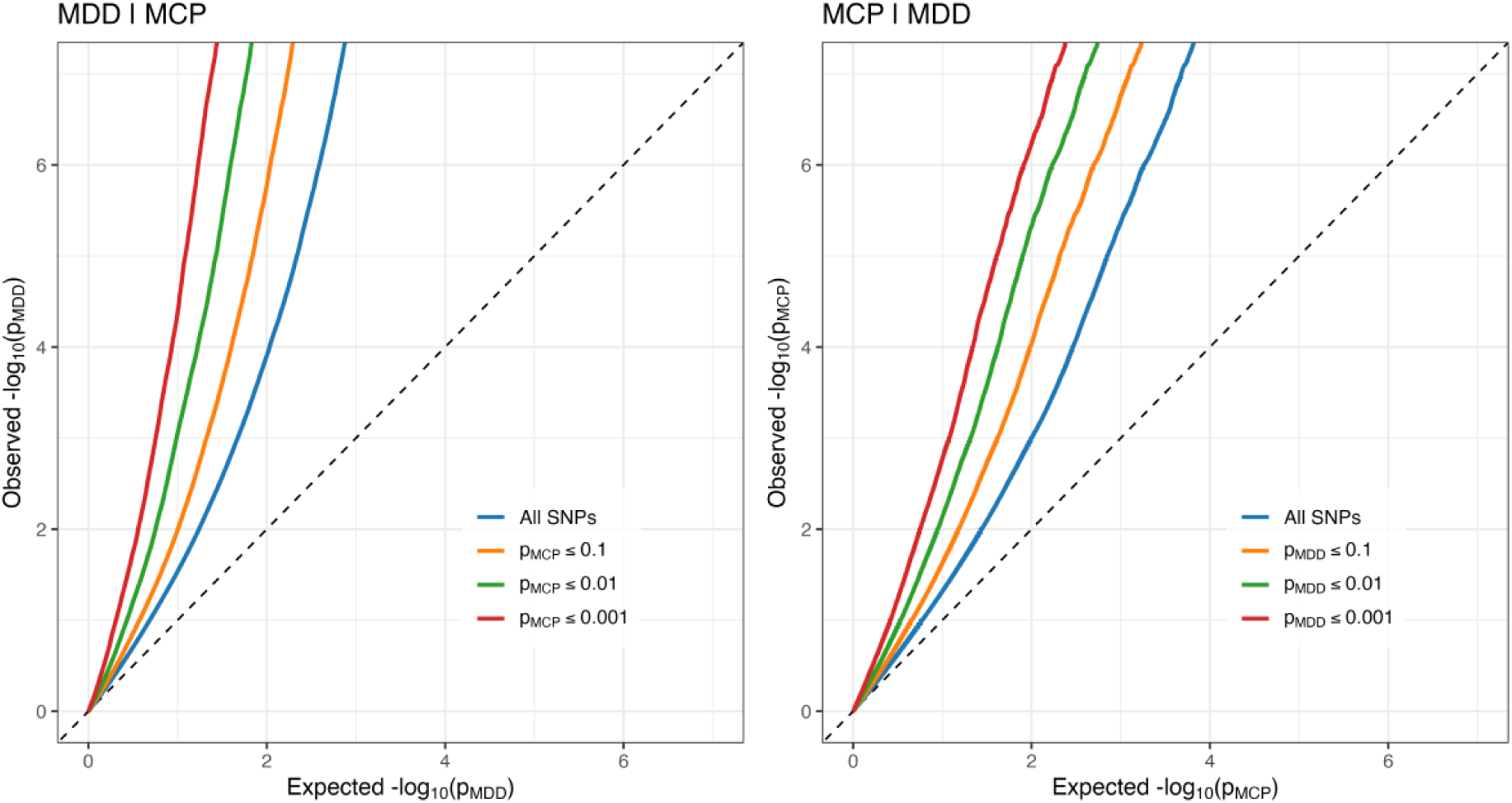
Polygenic overlapping effects between MDD and MCP. Conditional Q-Q plots of observed versus expected −log10(p) values: left, MDD conditional on MCP; right, MCP conditional on MDD. Curves show all genetic variants (blue) and variant subsets stratified by the other trait’s association strength (p ≤ 0.100, 0.010, and 0.001; orange/green/red). Upward deflection from the dashed diagonal (null expectation) indicates cross-trait enrichment consistent with polygenic overlap. MCP = multisite chronic pain, MDD = major depressive disorder.

### Shared genetic loci between MCP and MDD

The conjFDR analysis was used to identify shared loci between MCP and MDD. At a conjFDR < 0.05 threshold, we observed 483 lead SNPs across 375 independent genomic loci that were jointly associated with both MDD and MCP (Supplementary Table 3; Figure 3). Effect directions of shared lead SNPs were found to be 96.1% consistent, with 464 lead SNPs exhibiting the same effect direction in MDD and MCP and 19 showing opposite effect directions. Cross-trait genetic colocalisation was further applied to test whether MDD and MCP shared signals are driven by the same causal variant (PP4 > 0.8). Of the 375 shared genomic loci, 22 showed strong evidence of colocalisation (Table 1).

**Figure 3.**
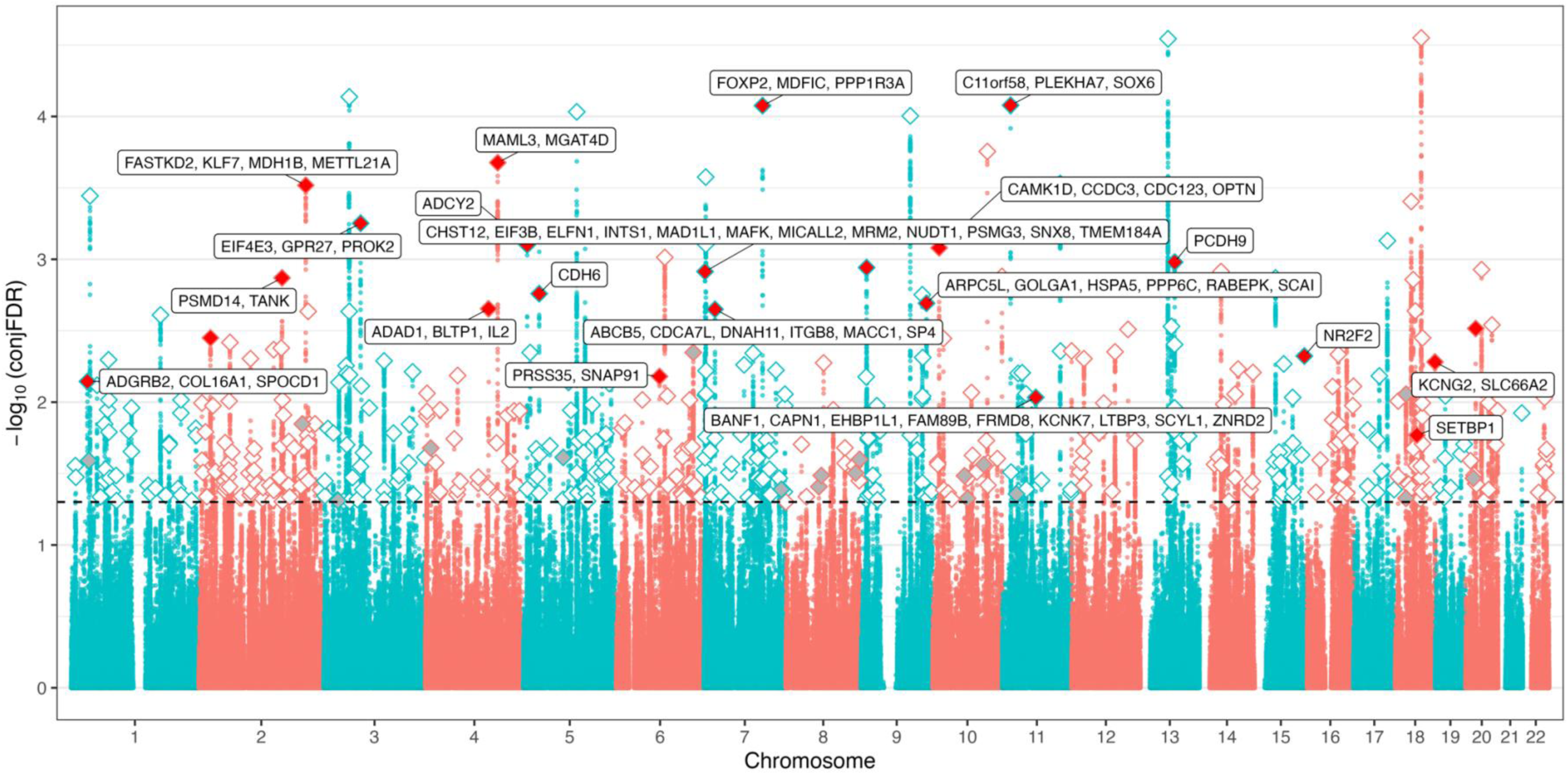
Shared genetic variants and genes between MDD and MCP. The Manhattan plot displays −log10(conjFDR) for each genetic variant on the y-axis against chromosomal position on the x-axis. The dashed horizontal line marks the significance threshold for shared associations (conjFDR < 0.05; i.e., −log10(conjFDR) > 1.3). White and grey diamonds represent lead SNPs with the same and opposite effect directions in MDD and MCP, respectively. Red diamonds represent lead SNPs at colocalised loci, all of which have consistent effect directions. Labels indicate the prioritised gene(s) at each colocalised locus as determined by locus-to-gene analysis

### Gene prioritisation and pathway enrichment

Across the 22 colocalised genomic loci, we mapped 69 associated genes using Open Targets’ L2G pipeline (Table 1, Figure 3). As of 30 Oct 2025, querying the Open Targets Platform (https://genetics-docs.opentargets.org/) indicated prior associations with depression- or pain-related conditions for all genes identified, with the exceptions of *family with sequence similarity 89 member B* (*FAM89B*), *Golgin subfamily A member 1* (*GOLGA1*), and s*olute carrier family 66 member 2* (*SLC66A2*). The 69 associated genes were enriched in the GO terms related to cadherin-mediated cell-cell adhesion, translational initiation, endosomal transport, Golgi vesicle transport, cellular response to transforming growth factor beta (TGF-β) stimulus and pregnancy and KEGG pathways of neurodegeneration and amyotrophic lateral sclerosis (ALS) (Supplementary Table 7).

**Table 1.**
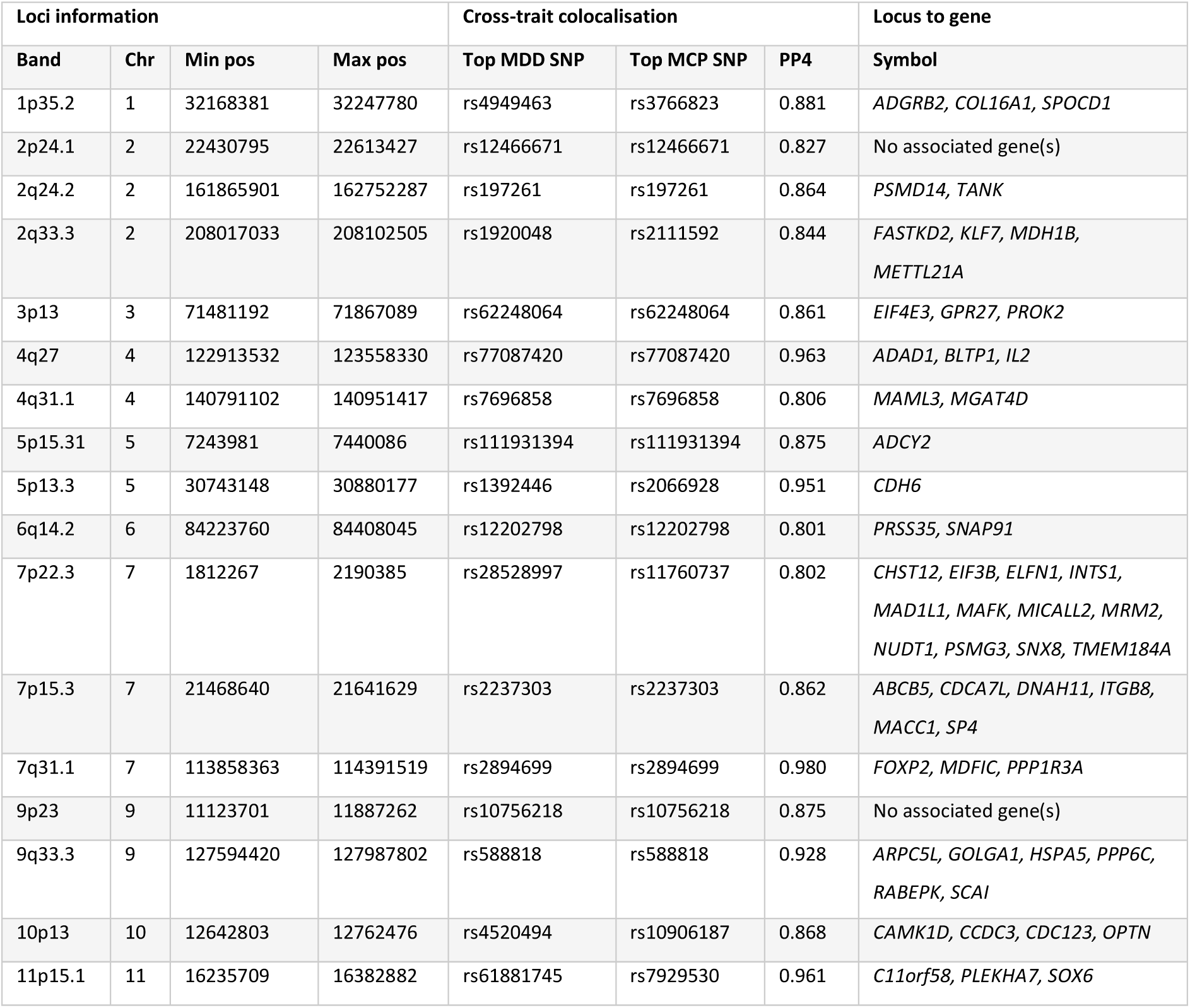
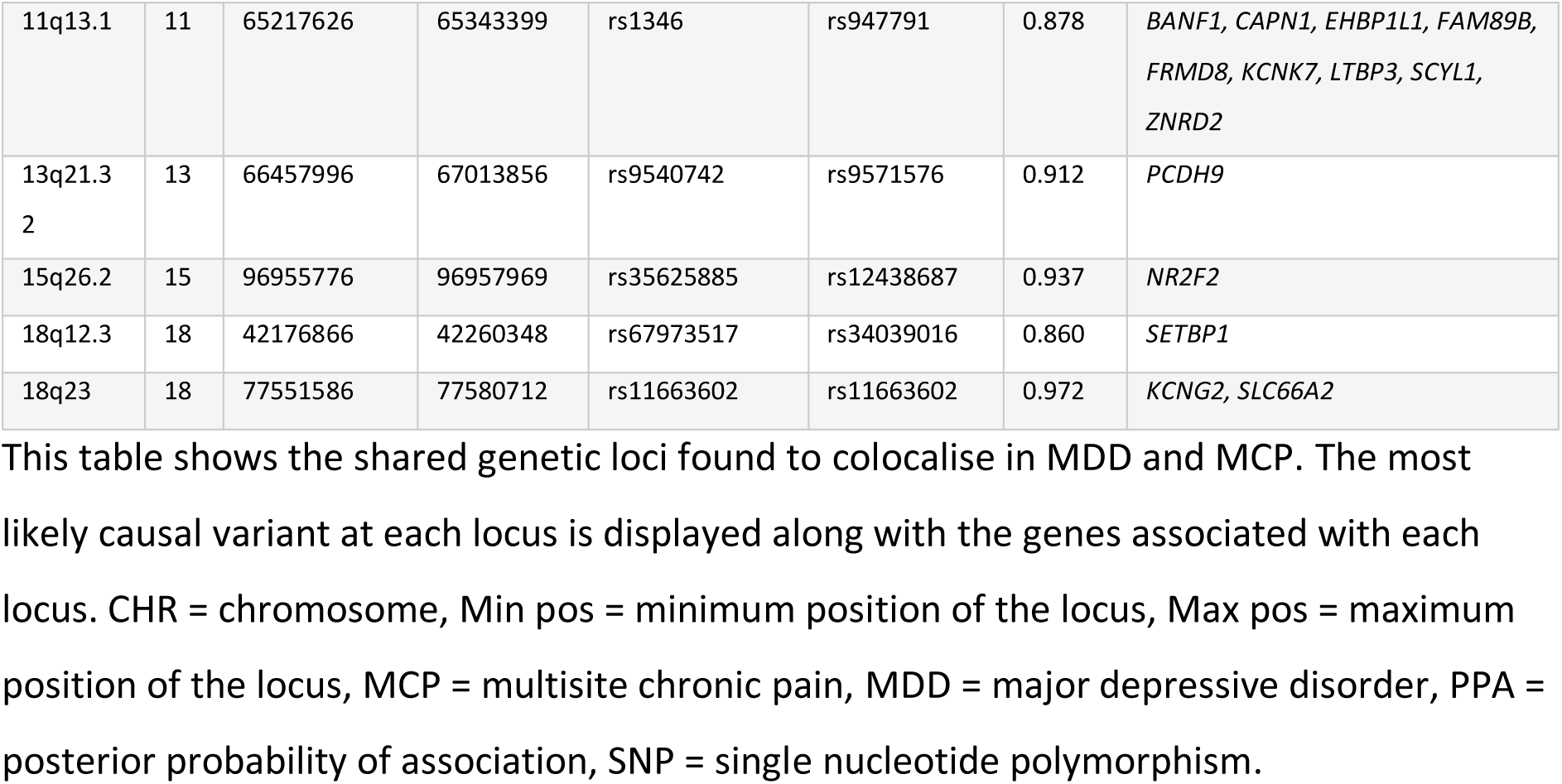
Colocalised loci of MDD and MCP.

### Shared colocalisation with gene expression in brain tissue

We performed genetic colocalisation tests to estimate the probability that the same single causal variant is causal for the expression of our prioritised genes in brain tissue and both MDD and MCP (Table 2). We found that both MDD and MCP had a shared colocalisation with *protein phosphatase 6 catalytic subunit* (*PPP6C*) in the caudate (MDD: PP4 = 0.920; MCP: PP4 = 0.919), cortex (MDD: PP4 = 0.900; MCP: PP4 = 0.814), frontal cortex (BA9) (MDD: PP4 = 0.909; MCP: PP4 = 0.893) and putamen (MDD: PP4 = 0.924; MCP: PP4 = 0.915) and *suppressor of cancer cell invasion* (*SCAI*) in the cortex (MDD: PP4 = 0.890; MCP: PP4 = 0.821).

Evidence for disorder-specific eQTL colocalisation in MDD was observed for the expression of *actin-related protein 2/3 complex subunit 5-like* (*ARPC5L*) in the hypothalamus (PP4 = 0.806), *G protein-coupled receptor 27* (*GPR27*) in the amygdala (PP4 = 0.829), caudate (PP4 = 0.867), cerebellar hemisphere (PP4 = 0.897), cerebellum (PP4 = 0.821), frontal cortex (BA9) (PP4 = 0.832) and hypothalamus (PP4 = 0.891), *PPP6C* in the hippocampus (PP4 = 0.827) and hypothalamus (PP4 = 0.839) and *Sp4 transcription factor (SP4*) in the nucleus accumbens (PP4 = 0.811). MCP was also specifically colocalised with *dynein axonemal heavy chain 11* (*DNAH11*) in the caudate (PP4 = 0.874) and *GPR27* in the nucleus accumbens (PP4 = 0.983).

**Table 2.** Brain tissue eQTL-GWAS colocalisation in MDD and MCP.

| GTEx Brain Tissue | Gene | PP4 MCP | PP4 MDD |
| --- | --- | --- | --- |
| Brain_Hypothalamus | *ARPC5L* | 0.663 | 0.806 |
| Brain_Caudate_basal_ganglia | *DNAH11* | 0.874 | 0.396 |
| Brain_Amygdala | *GPR27* | 0.768 | 0.829 |
| Brain_Caudate_basal_ganglia | *GPR27* | 0.781 | 0.867 |
| Brain_Cerebellar_Hemisphere | *GPR27* | 0.452 | 0.897 |
| Brain_Cerebellum | *GPR27* | 0.688 | 0.821 |
| Brain_Frontal_Cortex_BA9 | *GPR27* | 0.550 | 0.832 |
| Brain_Hypothalamus | *GPR27* | 0.298 | 0.891 |
| Brain_Nucleus_accumbens_basal_ganglia | *GPR27* | 0.983 | 0.000 |
| Brain_Caudate_basal_ganglia | *PPP6C* | 0.919 | 0.920 |
| Brain_Cortex | *PPP6C* | 0.814 | 0.900 |
| Brain_Frontal_Cortex_BA9 | *PPP6C* | 0.893 | 0.908 |
| Brain_Hippocampus | *PPP6C* | 0.697 | 0.827 |
| Brain_Hypothalamus | *PPP6C* | 0.719 | 0.839 |
| Brain_Putamen_basal_ganglia | *PPP6C* | 0.915 | 0.924 |
| Brain_Cortex | *SCAI* | 0.821 | 0.890 |
| Brain_Nucleus_accumbens_basal_ganglia | *SP4* | 0.729 | 0.811 |

This table shows the evidence of strong colocalisation between each trait (MDD and MCP) and cis-eQTLs of genes across GTEx brain tissues. *ARPC5L* = *actin-related protein 2/3 complex subunit 5-like*, *DNAH11* = *dynein axonemal heavy chain 11*, *GPR27* = *G protein-coupled receptor 27*, MCP = multisite chronic pain, MDD = major depressive disorder, PPA = posterior probability of association, *PPP6C* = *protein phosphatase 6 catalytic subunit*, *SCAI* = *suppressor of cancer cell invasion*, *SP4* = *Sp4 transcription factor*.

## Discussion

This study explored the shared genetic architecture between MDD and MCP using advanced analytical techniques and large-scale GWAS datasets. Our results from bivariate MiXeR analysis demonstrated that both traits have a high degree of directionally consistent polygenic overlap (Dice coefficient = 67.4%, concordant allelic effects = 89.8%). ConjFDR analysis identified 375 shared loci, 22 of which showed cross-trait colocalisation between the MDD and MCP signals. Gene mapping and enrichment implicated several biological processes, including cadherin-mediated cell-cell adhesion and translational initiation. Furthermore, gene expression colocalisation in brain tissue highlighted *PPP6C* and *SCAI* in both disorders.

Previous studies have quantified genome-wide genetic correlations between depression- and pain-related phenotypes using LDSC (Johnston et al., 2019a; Meng et al., 2020), which has been shown to underestimate the extent of genetic overlap in cases of mixed directions of effect across shared genetic loci (Frei et al., 2019). In contrast, bivariate MiXeR explicitly models the balance of effect directions among shared variants, making it sensitive to directionality, and can recover substantial polygenic overlap even when genome-wide genetic correlation is attenuated by mixed effects. Applied here, MiXeR provided novel insight into the extensive and largely consistent overlap between MDD and MCP, but was not shown to outperform LDSC as indicated by the negative AIC_best vs. min_ value obtained from the model. This is expected when most shared variants act in the same direction: under high concordance, LDSC captures the cross-trait signal well and MiXeR yields limited incremental gain (Frei et al., 2019).

Beyond global pleiotropy, conjFDR analysis identified 375 loci jointly associated with MDD and MCP, 22 of which showed strong cross-trait colocalisation (PP4 > 0.8). Prior work using the conjFDR framework with lower-powered GWASes of MDD and von Korff chronic pain grade (CPG) highlighted a single pleiotropic locus at *LRFN5* (Johnston et al., 2019b). Consistent with this, our results identified an intronic *LRFN5* variant, rs10138326 (Supplementary Table 3), as a shared lead SNP; however, there was no evidence of strong colocalisation at this locus (PP4 = 0.462), meaning we cannot distinguish this result as a product of a genuine shared causal variant or a result of LD. By leveraging cross-trait enrichment in the largest GWAS available for both traits, our conjFDR analysis has identified specific shared loci (with colocalisation support), extended prior applications of the method and provided novel, locus-level insight into the genetic aetiology common to MDD and MCP.

The 22 putative shared causal loci supported by cross-trait colocalisation mapped to 69 genes. Notably, three of these genes, *FAM89B*, *GOLGA1*, and *SLC66A2*, had no prior associations with depression- or pain-related phenotypes in the Open Targets Platform (accessed 30 Oct 2025). Gene-set enrichment analysis of all 69 genes elucidated their involvement in numerous biological processes: cadherin-mediated cell-cell adhesion, translational initiation, endosomal transport, Golgi vesicle transport, cellular response to TGF-β stimulus and female pregnancy. Although broad and heterogeneous, components of some of these pathways have previously been implicated in pain and mood. For example, cadherin-mediated adhesion supports the induction and maintenance of long-term potentiation (LTP) (Bozdagi et al., 2010), a process central to the development of chronic pain (Sandkühler, 2007) and implicated in depression-related behaviour (Liu et al., 2017; Yang et al., 2018); altered endosomal and Golgi vesicle trafficking changes the surface abundance and signalling of receptor and ion channels at synapses (Anggono & Huganir, 2012; Ferron et al., 2021; Lau & Zukin, 2007), thereby modulating synaptic strength and potentially disrupting pain and mood processing. Overall, these findings have identified specific genes and several putative biological pathways as potential targets for the development of novel therapeutic interventions for co-occurring chronic pain and depression.

Genetic colocalisation of eQTL data prioritised likely effector genes and brain tissues in MDD and MCP pathophysiology. Cis-eQTLs for *PPP6C* in caudate, cortex, frontal cortex and putamen colocalised with both traits (PP4 > 0.8). Effect directions at the colocalised, putative causal variants indicate that higher gene expression is associated with increased risk for both disorders (Supplementary Table 3; https://www.gtexportal.org). *PPP6C* has previously been identified in a brain transcriptome-wide association study (TWAS) of MCP (Johnston et al., 2024). *PPP6C* encodes the catalytic subunit of protein phosphatase-6 (PP6), a component of a signalling pathway regulating cell cycle progression, which has been demonstrated in preclinical studies to promote neurite outgrowth (Kitamura et al., 2021), and the development of cortical- and inter-neurons (Matsuoka et al., 2024). One possible explanation for our findings is that PP6-dependent regulation of neurite outgrowth and neuron development within cortico-striatal circuits (caudate/putamen and frontal cortex), which mediate sensory integration and emotion regulation (Borsook et al., 2010; Rive et al., 2013; Starr et al., 2011), contributes to the shared aetiology of MDD and MCP.

Genetically predicted expression of *SCAI* in the cortex was also found to colocalise with both MDD and MCP. Interestingly, *SCAI* sits in the same 9q33.3 locus as *PPP6C*, placing them within the same colocalised region. *SCAI* encodes a transcriptional cofactor, best known for its inhibition of the movement of cancer cells (Brandt et al., 2009). Based on the effect directions of putative causal variants, it is indicated that higher expression of *SCAI* is associated with increased risk for both disorders (Supplementary Table 3; https://www.gtexportal.org). Higher expression of *SCAI* has previously been associated with increased depression risk (Zeng et al., 2024), and toxic/inflammatory neuropathy (Johnston et al., 2024). Preclinical work has also demonstrated its role in neuronal differentiation, particularly dendritic morphology (Ishikawa et al., 2010). Further work investigating how cortical expression of *SCAI* might impact MDD and MCP pathophysiology is needed to fully understand these findings.

The proteins encoded by *PPP6C* and *SCAI* are not known drug targets. Moreover, both encoded proteins have tumour-suppressive roles. The PPP6C protein acts as a tumour-suppressive phosphatase and is frequently mutated or downregulated in melanoma (Cho et al., 2021). Similarly, the SCAI protein suppresses cancer cell invasion (Brandt et al., 2009). Therefore, systemic inhibition of these proteins raises safety concerns, as it could promote tumour progression, including invasion and metastasis.

Several additional gene expression-tissue pairs colocalised with only one disorder. *GPR27* expression colocalised with MDD in the amygdala, caudate, cerebellar hemisphere, cerebellum, frontal cortex and hypothalamus, and with MCP in the nucleus accumbens, implicating the same gene in both disorders but in non-overlapping tissues. Other disorder-specific colocalisation included *ARPC5L* expression in the hypothalamus (MDD), *SP4* expression in the nucleus accumbens (MDD) and *DNAH11* in the caudate (MDD). These findings warrant further investigation to clarify mechanism and therapeutic potential in each disorder.

One major strength of this study is the use of advanced genetic analysis techniques and large, well-powered GWAS datasets to gain robust insights into the pleiotropy between MDD and MCP, on both a global and locus-specific level. By anchoring biological inferences for MDD and MCP, two very complex disorders, in shared loci, we also minimise reverse-causation concerns: because genotypes are fixed (Davey Smith & Hemani, 2014), the implicated pathways are more likely to reflect common causal mechanisms rather than downstream consequences of disease.

When interpreting the findings of this study, it is also important to acknowledge several important limitations. First, in the absence of adequately powered and independent multi-ancestry GWAS, our analysis was restricted to individuals of European ancestry, limiting the generalisability of our findings. Second, the GWAS inputs were restricted to common autosomal variants, so shared risk mediated by rare variants was not assessed. Finally, the contribution of mediated (vertical) pleiotropy to the observed genetic overlap is unclear, making it hard to distinguish shared biology from genetic liability that first affects one trait and thereby increases risk of the other.

## Conclusion

Using innovative genetic approaches, this study showed extensive, largely concordant polygenic overlap between chronic pain and depression and pinpointed loci colocalised across both traits. Functional annotation implicated biological pathways central to synaptic plasticity and signalling, and eQTL colocalisation highlighted *PPP6C* and *SCAI* expression in brain tissues as mediators of shared risk. Overall, these findings have enhanced our understanding of the complex association between chronic pain and depression by identifying shared molecular mechanisms that warrant further study as novel targets for prevention and treatment.

## Data availability

All GWAS summary statistics used in this study are publicly available. Specifically, MDD summary statistics can be downloaded from https://pgc.unc.edu/for-researchers/download-results/, and MCP summary statistics are available at https://researchdata.gla.ac.uk/822/.

## Code availability

The custom code used to perform all analyses for this study will be made publicly available after publication at https://github.com/hannahcasey/MDD_MCP_pleiotropy.

## Author contributions

**Hannah Casey:** Conceptualisation; data curation; formal analysis; methodology; software; validation; visualisation; writing - original draft; writing - review & editing

**Xueyi Shen:** Conceptualisation; methodology

**Laurence Nisbet:** Conceptualisation; methodology

**Marie T. Fallon:** Conceptualisation; supervision; writing - review & editing

**Daniel J. Smith:** Conceptualisation; supervision; writing - review & editing

**Rona J. Strawbridge:** Conceptualisation; methodology; supervision; writing - review & editing

**Heather C. Whalley:** Conceptualisation; methodology; supervision; writing - review & editing

